# Textile Masks and Surface Covers – A ‘Universal Droplet Reduction Model’ Against Respiratory Pandemics

**DOI:** 10.1101/2020.04.07.20045617

**Authors:** Alex Rodriguez-Palacios, Fabio Cominelli, Abigail R. Basson, Theresa T. Pizarro, Sanja Ilic

**Affiliations:** Division of Gastroenterology & Liver Diseases, Case Western Reserve University School of Medicine; Digestive Health Research Institute, University Hospitals Cleveland Medical Center, Cleveland, OH, USA; Department of Pathology, Case Western Reserve University School of Medicine; Human Nutrition, Department of Human Sciences, College of Education and Human Ecology, The Ohio State University, Columbus, OH, USA

**Author notes:** **Corresponding Author:** Alex Rodriguez-Palacios.

**Keywords:** Coronavirus, respiratory pandemic, COVID-19, SARS-Cov-2, Cloth Masks, Fabrics

## Abstract

The main form of COVID-19 transmission is via ‘oral-respiratory droplet contamination’ (droplet; very small drop of liquid) produced when individuals talk, sneeze or cough. In hospitals, health-care workers wear facemasks as a minimum medical ‘*droplet precaution*’ to protect themselves. Due to the shortage of masks during the pandemic, priority is given to hospitals for their distribution. As a result, the availability/use of medical masks is discouraged for the public. However, given that asymptomatic individuals, not wearing masks within the public, can be highly contagious for COVID-19, prevention of ‘*environmental droplet contamination*’ (EnDC) from coughing/sneezing/speech is fundamental to reducing transmission. As an immediate solution to promote ‘*public droplet safety’*, we assessed household textiles to quantify their potential as effective environmental droplet barriers (EDBs). The synchronized implementation of a universal ‘*community droplet reduction solution*’ is discussed as a model against COVID-19. Using a bacterial-suspension spray simulation model of droplet ejection (mimicking a sneeze), we quantified the extent by which widely available clothing fabrics reduce the dispersion of droplets onto surfaces within 1.8m, the minimum distance recommended for COVID-19 ‘social distancing’. All textiles reduced the number of droplets reaching surfaces, restricting their dispersion to <30cm, when used as single layers. When used as double-layers, textiles were as effective as medical mask/surgical-cloth materials, reducing droplet dispersion to <10cm, and the area of circumferential contamination to ∼0.3%. The synchronized implementation of EDBs as a ‘community droplet reduction solution’ (*i*.*e*., face covers/scarfs/masks & surface covers) could reduce EnDC and the risk of transmitting or acquiring infectious respiratory pathogens, including COVID-19.

## INTRODUCTION

The main form of COVID-19 transmission is via ‘*oral-respiratory droplets’* produced when individuals talk, sneeze or cough. Despite the magnitude of the COVID-19 pandemic, it is disconcerting that the general public, either does not have personal protective equipment available to them, including respiratory masks, or chose to not use them, to contain the pandemic.

Worldwide, health-care workers wear medical masks as a minimum ‘*droplet precaution*’ to protect themselves. However, experts appealed to the community not to wear medical masks stating they are not effective for the public^1^; albeit, counter-criticisms ensued^2^. Regardless of clinical presentation, COVID-19 transmits person-to-person, including children^3^ via ‘oral-respiratory droplets’ produced when individuals talk or sneeze/cough. Aside from Asia^4^, there are no global guidelines promoting wearing masks in public to control respiratory pandemics^5-10^, and no scientific data/guidelines exist promoting masks as a ‘droplet precaution’ for the public^5,9,11^

COVID-19 is caused by a novel coronavirus strain (SARS-CoV-2), for which there is no treatment^12,13^. Disease is characterized by fever, coughing/sneezing, dyspnea, pneumonia, and death^14^; however, important for asymptomatic transmission, cases increasingly present with gastrointestinal symptoms, and/or fatigue, without fever^15^. Regardless of the clinical presentation, COVID-19 transmits person-to-person through oral-respiratory droplets produced when infected individuals (symptomatic or asymptomatic, including children^3^) talk/cough/sneeze, contaminating the environment.

Although viruses can become airborne dust/aerosols, as micro-droplets evaporate, viruses rapidly loose infectivity in the air (half-life=1h)^16-20^. By contrast, virus survival increases when liquid droplets contaminate surfaces, especially, plastic and stainless steel, with long half-lives of 7 and 6h, respectively (cardboard, 4h; copper, 1h)^16^. Since COVID-19 transmits when droplets reach nose/mouth/eyes^21^, or when people touch their nose/mouth/eyes after touching droplet-contaminated surfaces (supermarkets/elevators^22^), it is critical to implement strategies to prevent/reduce environmental droplet contamination (EnvDC). Particularly, because plastic and metal surfaces remain infective for days. Herein, we investigated whether common household textiles can be used as environmental droplet barriers (EDBs; facemasks/covers/scarfs, or surface covers) to prevent EnvDC, improving public droplet safety, and support the synchronized implementation of an environmentally-purposed *Universal Droplet Reduction Model* within the public to control COVID-19.

## METHODS

### Simulation of bacteria-containing micro-/macro-droplet clouds

Since viruses exist in association with bacteria and host cells within electrolytes-rich respiratory fluids ^23,24^, we used a bacterial-suspension strategy to quantify the number of droplets that could not be visualized, but that could escape textile barriers and cause long-/short-range surface contamination. To enumerate bacteria-carrying micro-droplets, we used household spray bottles filled with an aqueous suspension of 12-probiotic-cultured dairy product (*Lactobacillus lactis, L. rhamnosus, L. plantarum, L. casei, L. acidophilus, Leuconostoc cremoris, Bifidobacterium longum, B. breve, B. lactis, Streptococcus diacetylactis*, and *Saccharomyces florentinus*, 75ml; 3×10^6-7^ cfu/ml) in 1000ml PBS (Fisher BP-399-1) to simulate a cloud of droplets produced by a sneeze. Probiotics are BSL-1/’Generally Recognized As Safe’ by the FDA and all experiments were conducted in BSL-2 HEPA-filtered microbiology laboratories. No animal/human subjects were used for experimentation. Before testing, spray bottle nozzles were adjusted to produce cloud and jet-propelled droplets that match the visual architecture of droplet formation described by Bourouiba *el at*.^23^. Quantification of droplets landing over a surface was performed at the time of spray using seven 10mm-Petri dishes containing tryptic soy agar (56.75cm^2^ surface area/dish) with 5% defibrinated sheep blood, placed on a table spaced at 30cm intervals between 0-180cm. Plates remained open for 10 minutes to allow droplet landing. Droplet quantification was conducted for each bottle in duplicate. Large-drop quantification outside agar plates was facilitated by a white droplet footprint left on black surfaces.

### Quantification of droplet retention by household textiles

To simulate the function of mask barriers, we placed selected textiles (∼22X22cm) over a cardboard/plastic-covered 25X30cm surface, over a carved (8.5×11cm) window, at 8.5cm from the agar plates’ plane, through which droplets were sprayed. On the other side of the textile, 3-5 agar plates were aligned to cover the 0-8.5, 8.5-17, 17-25.5, 25.5-34cm intervals to quantify bacteria-containing droplets that could contaminate a surface. Quantification represents droplets that pass through the textile and that land on a rectangular area of 8.5cmX180cm (agar plate diameter X ‘spray path’). To quantify the effect of textiles retaining vertically-landing droplets, we quantified droplets reaching agar plates covered with a household textile.

### Household textiles tested, replication of findings, and safety

We first tested six household textiles, including 100% combed cotton (widely available, ‘T-shirt material’), 100% polyester microfiber 300-thread count fabric (pillow case), two loosely woven ‘homespun’ 100% cotton fabrics (140GSM, 60×60-thread count; and 115GSM, 52×48-thread count), and ‘dry technology’ 100% polyester common in sport jerseys. These textiles were compared to: ***i)*** the lack of textile barrier (no mask control), ***ii)*** medical masks, and ***iii)*** surgical cloth material as ‘gold standard’ protective controls. To ensure external validity/reproducibility, complementary and repeated experiments were conducted with selected textiles (*i*.*e*., respiratory mask, sports jersey, and Cotton-T-shirt) at The Ohio State University.

### Statistical Analysis

Student’s-T tests, linear regression, and multinomial logistic regression were conducted using raw and Log_2_ transformed CFU data (STATA, v15.1). Confidence intervals are provided to convey information relevant to sample size and external validity.

## RESULTS

### Spray dispersion model of droplets reach >1.8 meters if upward

Because viruses replicate in bodily fluids, in association with other microorganisms^23,24^, and need droplets to facilitate their expulsion, transmission and EnDC^12^, we first validated a rapid spray-simulation model of droplets (mimicking a sneeze) using a bacterial-suspension to quantify the extent by which widely-available household textiles reduced the ejection/long-distance flight of droplets. To facilitate the enumeration of macro-droplets and invisible micro-droplets, spray-simulations were conducted over nutritious-media agar surfaces, incubated for 24h to enable colony-forming-droplet-unit (CFDU) formation.

Based on simulations conducted in two institutions, a cloud of bacteria-carrying droplets travel distances reaching >180cm, particularly for large droplets (**Figure 1A**), consistent with reported dynamics during sneezing^23^. Of relevance to sneezing behavior, simulations illustrate that upward inclination of the central-spray angle allows macro-droplets to reach longer distances (simulation 4/dispersion equations; **Figure 1B-E**). Although macro-droplets frequently reached 180cm, most micro-droplets landed on surfaces within 120cm, with spray air-turbulence carrying micro-droplets into areas not reached due to gravity alone. Thus, social distancing of 1.8m, without EDB-mask protection, as currently recommended, is insufficient to prevent droplet exposure, particularly where essential-service workers congregate during pandemics (transportation, supermarkets/food displays). Therein, wearing EDB-masks together with inclining downward the head/body during sneezing could minimize the spatial range of EnDC.

**Figure 1.**
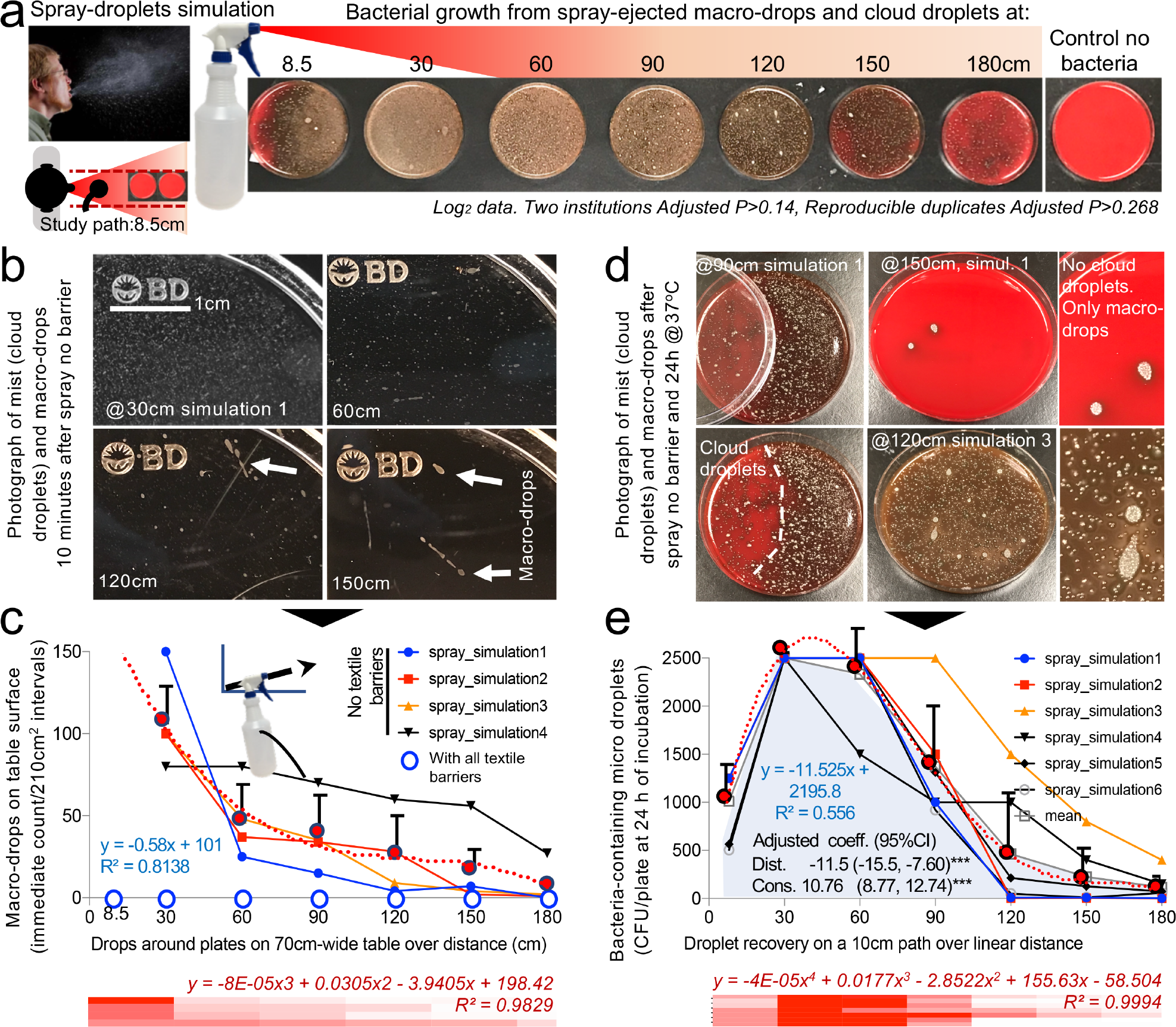
Simulation of a cloud of airborne bacteria-containing macro-drops and micro-droplets to quantify barrier potential of household textiles. **A)** Graphical overview of spray model as shown in Figure 1A. Inset, Photograph of a human sneeze, public domain, James Gathany, CDC image ID11162). **B**) Photographs of short and long-range visible droplets immediately after spray. **C**) Number of macro-drops for four simulations over distance. **D**) Photographs of bacterial CFDUs on agar plates illustrating ability of cloud micro-droplets to move around spaces driven by cloud turbulence (left images, agar plates were partially covered with lid at moment of spray), concurrent contamination with macro- and micro-droplets. **E**) Number of CFDU/plate (56.75 cm^2^) for 6 simulations over distance.

### Household textiles retain liquid droplets, particularly if double layered

To quantify the droplet retention potential of textiles as EDBs, we next used the same bacterial-spray-simulation model to quantify non-visualizable micro-droplets that could cross/escape the textile-EDB and cause microbial-surface agar contamination (textile/thread details in **Supplementary Figure 1**). Textiles were tested for one- and three-sprays to determine if EnDC changed with textile humidity. Although humidity had no statistical impact (dry-vs-humid, adj.-P>0.2), all textiles, tested as ‘single-layers’, significantly and reproducibly (between institutions) reduced the ejection of macro-droplets, and the traffic of micro-droplets to <25.5-34cm (linear regression adj.-P<0.001, compared to 180cm with no textile barrier; **Figure 2A-B** and **Supplementary Figures 2-3**).

**Figure 2.**
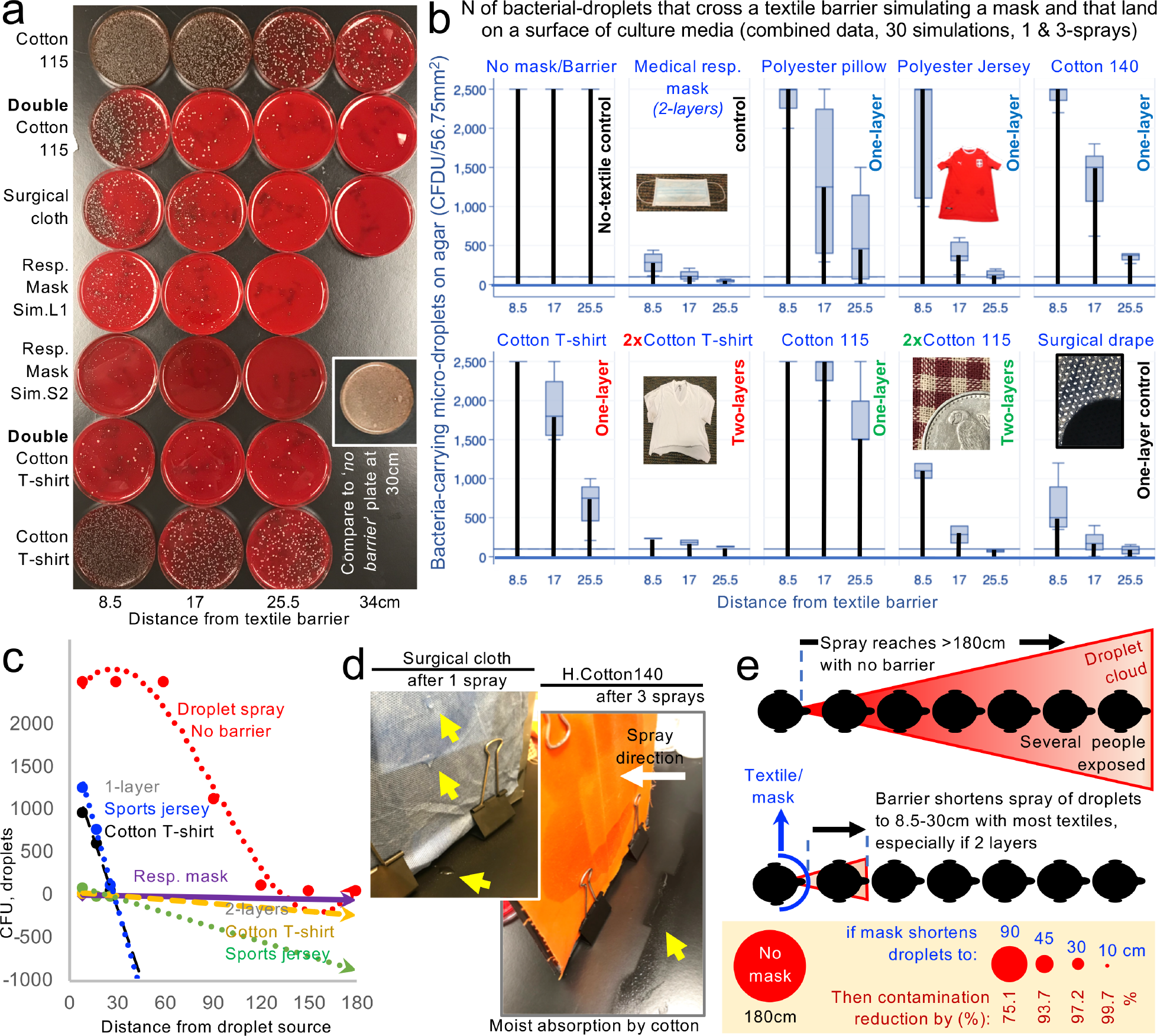
Using two layers of household textiles markedly retain liquid droplets. **A**) Tryptic soy agar plates illustrate effective bacterial-droplet reduction by 2-layer textiles. **B**) Pooled results from two spray-simulations (1- and 3-sprays; **Supplementary Figure 2**). **C**) Linear regressions for EnDC reduction over distance for no-barrier vs. selected textiles. Compared no textile (EDB) barrier (red dotted line), the reduction in CFUs illustrate the profound effect of using household textiles to retain droplets. Line slopes that are closer to the horizontal grid line at 0, and closer to the ‘Resp. mask’-dotted line are more effective strategies (commercial masks are made of 2-or-3-layers) compared to single layers. (**Supplementary Figure 4**, equations and R^2^). **D**) Photographs of differences in moisture retention. Arrowheads, drops/accumulation. **E**) Benefit of wearing textile-EDBs in reducing circumferential EnDC.

Remarkably, spray experiments with ‘two-layers’ (of 100%-combed cotton, common in t-shirts; and 100% polyester, in sports jerseys) completely prevented the ejection of large macro-droplets (100% EnDC prevention), and drastically reduced the ejection of micro-droplets by a factor of 5.16Log2, which is equivalent to a 97.2% droplet reduction (P<0.020 vs. single-layers, **Figure 2C** and **Supplementary Figures 4-5**). Importantly, the least-effective textile as single-layer (most-’breathable’, 100%-cotton homespun-115 material) achieved a 90-99.998% droplet retention improvement when used as two-layers (95%CI=3.74-15.39Log2). Lastly, all textiles were equally effective at absorbing the humidity from 3-sprays compared to medical mask/surgical cloth materials, which condensate after 1-spray (**Figure 2D**). Together, experiments indicate that two-layers of household textiles are as effective as medical masks preventing EnDC, and that more breathable materials in ≥2-layers could be effectively used if individuals deem two-layer, ‘denser’ textiles too air-restrictive.

### ‘Universal Droplet Reduction Model’ against Rapid Respiratory Pandemics

We then rationalized the potential impact of a ‘universal droplet reduction model’ against pandemics, where the community act together to miniaturize the spatial range of EnDC. Since it is unclear how many viral particles in droplets (virus/μm^3^) or surfaces (virus/cm^2^) are needed to acquire COVID-19, we assumed that any droplet on a surface area of 56.75cm^2^ (an 8.5-cm diameter agar plate) renders a surface infective. Since textiles prevented droplets from reaching beyond a ∼30-cm radius, we propose a working ‘droplet reduction model’ to control COVID-19, where EDB-masks could reduce the ‘circumferential area of contamination’ around each individual by 97.2% when used as single-layers, or as much as 99.7% when used as two-layers. Especially 100%-cotton/polyester which shortened the EnDC radius to <10cm (similar to medical-mask material; Log_2_ difference=0.06, for 100% polyester, multinomial adj.-P>0.6). Because COVID-19 cases increase daily, and the fabrication of EDB by centralized organizations could take weeks to reach entire ‘lockdown’ communities, we suggest, based on the cotton/polyester EnDC effectiveness, and a homemade EDB-mask fabrication trial (**Supplementary Figure 6**), that, from one piece of clothing, every individual could make (without sewing machine) two 2-layer-EDB masks as an immediate, synchronized contribution to reduce COVID-19 EnDC.

From a surface perspective, if everyone were encouraged to wear EDBs, the collective area contaminated with droplets would be miniaturized to 0.3-2.77% (two-layers/single-layers), compared to the potential contamination within 180cm (10.2 m^2^). Even suboptimal EDBs, effective for 90cm radius, could mathematically reduce the EnDC area by 75.1% (**Figure 2E**). Our findings and surface estimations are conservative as they are based on simulations using a (non-viscous) liquid solution, assuming stationary individuals. However, the impact of EDB is predictably greater since real/large viscous secretions (**Figure 3A**), which also travel long distances (>180cm)^23^ would be easier to contain by EDB, as communities mobilize. To further lower the risk of fomite (plastic/metal surface) transmission from/by non-EDB-wearers, EDB-textiles used as covers, when relevant, could reduce EnDC by 90-98% (T-test P=0.003, **Figure 3B**).

**Figure 3.**
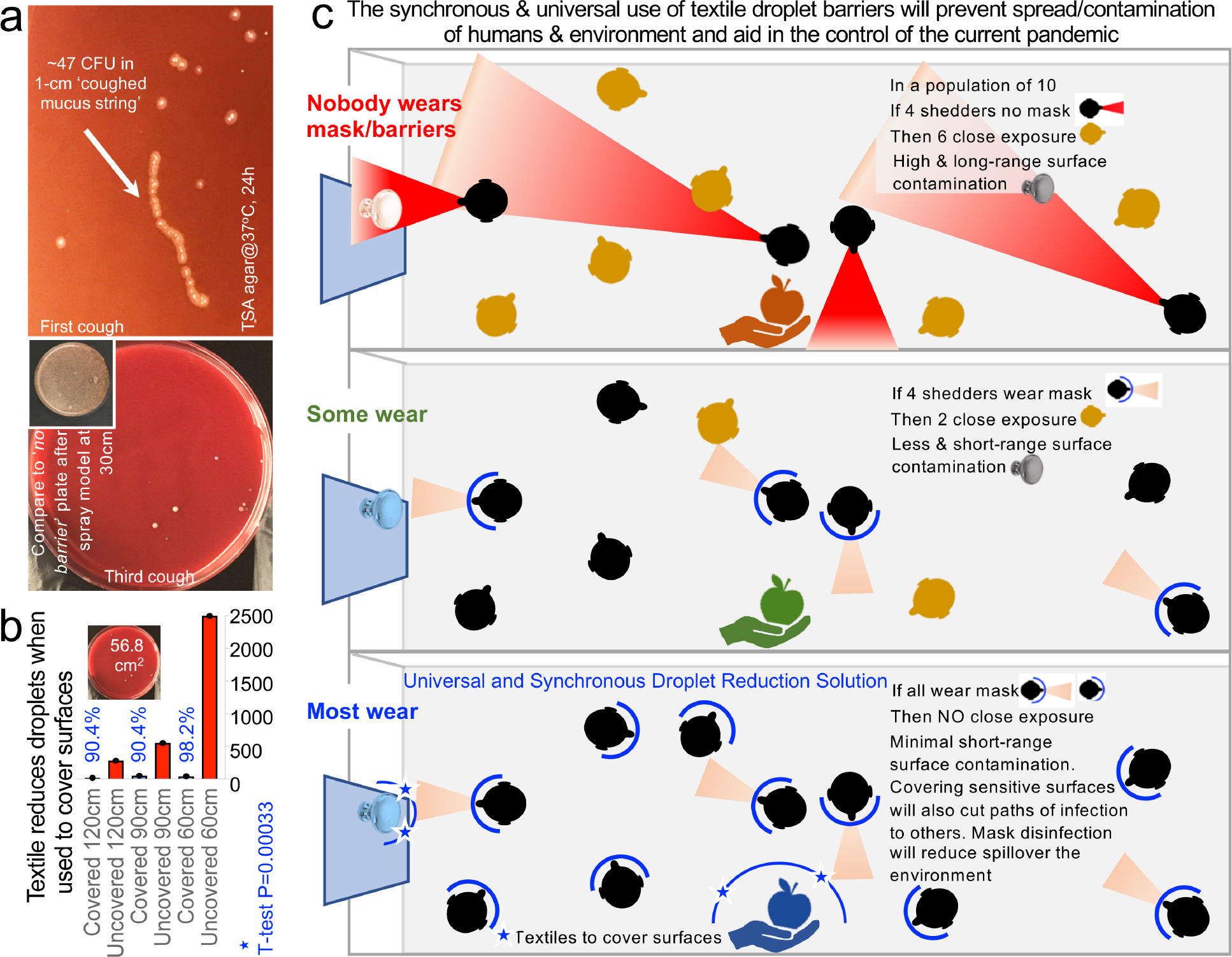
Environmentally-focused ‘Universal Droplet Reduction Model’ against pandemics due to infectious agents transmitted via oral-respiratory fluids. **A**) Coughed material-associated bacteria in agar. Large viscous secretions will be retained by textile-EDB. **B**) Bacteria-carrying droplet counts on agar plates covered with 1-layer cotton t-shirt material, after one-spray, over distance. **C)** Environmental droplet reduction model. Protective masks and surface covers in the community. **Supplementary Table 2**, list of current and proposed actions against COVID-19.

Finally, to illustrate, in volumetric terms, that EDBs are even more effective preventing EnDC, we conducted a scoping review of literature to conduct analyses of droplet fluid-carrying capacity. Although published droplet sizes vary with study method (**Supplementary Table 1**), most sneezed droplets are ‘large’, and can reach >1mm. Physiologically, two types of sneeze exist^25^: unimodal, when all droplets are large (360±1.5μm-diameter); and bimodal, when droplets are large (390±1.7μm-diameter, 70%) and small (72±1.5μm, 30%). Assuming droplets are spherical, for an average of two sneezes (unimodal:bimodal, 200,000 droplets), we determined that large droplets (85% of total) contain 703-times more fluid than small droplets. Thus, EDBs could profoundly reduce COVID-19 EnDC by effectively blocking the dispersion of fluids/viruses contained in large droplets. Because droplets of <47μm are known to evaporate before reaching the ground^26^, EDB will also prevent small-size droplet aerosolization by trapping such droplets immediately after production. An overview of a ‘universal textile droplet reduction action-model’ against pandemics is illustrated in **Figure 3C**.

## DISCUSSION

Despite widespread dissemination of information to curtail the rapid spread of COVID-19 outside of China (which affects 20-54-year-old adults, 40% of hospitalizations in the USA^27^), little attention has been devoted to EnDC and prevention strategies for droplet movement from infected to non-infected individuals within the same community. More concerningly, is that following mandatory ‘stay-in-home’ quarantine orders, people may return to work unprotected, unaware if they are infected/shedders. This is particularly critical for ‘essential pandemic workers’, who face different levels of risk (health-care vs. electric/transport/food services), and who can contaminate environmental surfaces as they transit through the community between work (*i*.*e*., hospitals) and home, or within their households^28^, without wearing masks. Because mass testing is not always possible^6^, especially for novel organisms like COVID-19, there are growing concerns that asymptomatic and mildly symptomatic citizens will continue to spread and reintroduce the virus to new areas, creating waves of cases, contributing to further economic burden from the outbreak^29^.

Nonpharmaceutical interventions (NPIs), also known as community mitigation strategies, are actions that individuals and communities can take in order to slow the spread of illnesses. For pandemics, when medical approaches (hospitalization/treatments) are limited, NPIs are a critical component to achieve resolution. Although PPE, including masks, are scientifically-effective to prevent infectious disease transmission, the use of masks for the general public has not been encouraged by governments^5,7^, possibly because demand will deepen the current crisis of mask unavailability for medical staff, or alternatively, because the use of masks to prevent respiratory infections has been misleadingly deemed ineffective, despite earlier clinical studies indicating that masks could be beneficial in households during pandemics^28,30,31^.

Although masks have been extensively studied to determine whether individuals are ***clinically*** protected from infections^32,33^, and to confirm that wearing a mask promotes desirable hygiene practices (handwashing, ‘avoiding crowds’)^5,31,34^, masks have not been examined for their potential to ***prevent environmental contamination***. Masks work, if worn properly; however, individuals (∼50%) often fail to wear masks regularly, and properly^30,35^. Despite low compliance, meta-analyses indicate that masks lower the odds of having (SARS)-respiratory infections by 87% (OR=0.13), compared to the odds of having an infection ‘not wearing a mask’^36^.

Herein, we propose, that in addition to seeking the classical/clinical ‘*prevention of infection’*, NPIs could be universally based on ‘*droplet reduction models*’ such as EDB to mitigate EnDC. Not only for the prevention of respiratory diseases, but also to prevent widespread environmental dispersion of the virus, which could reach water sources or affect domestic animals, as has been shown for other viruses, including pandemic influenza^37^.

The world is in short supply of masks since the international ‘lockdown’ affected production^38^, with health-care workers experiencing high morbidity/mortality due to reduced protection^39^. Governments are seeking private support to increase mask supplies; however, such strategy could take weeks/months, and infection rates would not improve if supplies were still not available to ‘lockdown’ communities. Increased community transmission leads to higher demand for medical services, unless transmission is halted. Using household textiles is a potentially life-saving cost-effective anti-pandemic strategy because washing/laundering textiles [destroys COVID-19 by heat (70°C/5min), bleach (1:49/5min), detergents (20min)], and is more sustainable (community-level) than using scarce medical disinfectants/supplies.

In conclusion, we demonstrated that, two-layer household textiles produced a profound reduction of EnDC as effectively as medical-grade materials. Encouraging/mandating the synchronous implementation of textile-EDBs, while discouraging using medical masks in public, could help control COVID-19.

## Data Availability

All data are available in the supplementary information files.

https://figshare.com/s/f9fc1f269a7406eed908

## Availability of data and materials

The raw data supporting the conclusions of this article will be made available by the authors, without undue reservation.

## Author contributions

ARP envisioned, planned and executed the experiments, analyzed the data, prepared figures and wrote the manuscript. SI executed validation and complementary experiments, interpreted data and wrote manuscript. ARB, assisted with documentation in Supplementary materials, commented and edit the paper. FC commented, revised and edited the manuscript for medical accuracy and data interpretation. All authors approved the final manuscript.

## Acknowledgements

NA.

